# Outbreak investigation of Visceral Leishmaniasis in Borena Zone, Oromia Region, Ethiopia, November 2019: Case Control study

**DOI:** 10.1101/2020.11.02.20222869

**Authors:** Belay Regassa, Negussie Deyessa, Adamu Addissie, Abdulnasir Abagero, Gemechu Shumi, Gemechu Gudina

**Affiliations:** Department of Public Health Emergency Management, Oromia Regional Health Bureau, Addis Ababa, Ethiopia; Department of Preventive Medicine, School of Public Health, College of Health Sciences, Addis Ababa University, Addis Ababa, Ethiopia

**Keywords:** Borena, Outbreak, Visceral Leishmaniasis

## Abstract

**Background:** Visceral Leishmaniasis (VL) caused by *Leishmania* parasites, infects mammals transmitted by *Phlebotomine* sand-flies and mostly affects the poorest. VL distributed worldwide and prevalent in Ethiopia. Knowing occurrence of disease and risk factor is a remedy for controlling. The aim of study was to identify factors associated with VL.

**Methods:** Case control study was carried out during October-November 2019 in Borena. A 1:2 Cases and controls were identified by case definition and 33 cases were included in the study. Participants >18years interviewed and caregivers of <18 were questioned for legal issue. Epi-info and SPSS were used for data entry and analysis. Primarily predictors were identified using chi-square at significant level P<0.05 with 95%CI, then candidate predictors were analysed using bivariate and multivariate analysis to identify associated factors.

**Result:** Among 153 suspected cases, 9 suspected deaths reported; 33 (22%) cases and 3 deaths were verified for VL. Among 33 verified cases 15(45.5%) were in July 2019, in comparison of 4years data, there is surge cases in July-August 2019, 26(79%) of cases were from Dire, Attack Rate (AR) = 15/100,000, CFR=9.1%. Among all, 15-64year were highly affected with AR=19.3. A case control engaged 99(100%) respondents and among all 93(93.9%) were male, 68(68.8%) were 15-64years. Adult education Adjusted Odds Ratio (AOR) = 30.438(2.378, 389.602), bed-net AOR=9.024 (1.763, 46.205) and walling AOR=0.052(0.004, 0.739) were associated factors with VL at 95%CI with p-value<0.05.

**Conclusion:** Male 15-64years were highly susceptible. Level of education, ITNs and walling were associated factors with VL. Formulating policies and guidelines for male 15-64 years related vector control and awareness creation regarding feeding habit of sand fly, prevention and control were recommended. Awareness of community on prevention method; using repellents, ITNs utilization, and safe sleeping mechanisms are mandatory. Further investigation on the issue is best remedy to overcome future VL outbreak occurrence.

## Introduction

Leishmaniasis is a protozoan disease caused by members of the genus *Leishmania*, parasites that infect numerous mammal species including humans, and transmitted by *Phlebotomine* sand-flies (1; 2; 3). Leishmania species produce widely varying clinical syndromes ranging from self-healing cutaneous ulcers to fatal visceral disease with the syndromes fall into visceral leishmaniasis (VL), cutaneous leishmaniasis, and mucosal leishmaniasis (3). Common symptoms of VL are prolonged fever, weight loss, signs of bone marrow invasion (anaemia, thrombocytopenia and leukopenia), abdominal distension with hepatosplenomegaly, and lymphadenopathy (4).

Transmission may be anthroponomical or zoonotic. Human-to-human transmission via shared infected needles and in-utero transmission to the foetus occurs rarely (3). Although the distribution of Leishmania is limited by the distribution of sand-fly vectors, human leishmaniasis is on the increase worldwide (2; 3). It affects the poorest and most marginalized people and is commonly associated with malnutrition, poor housing and weak immune system (5). Kala–azar generally affects poor and neglected populations living in remote rural areas. If not treated, more than 95% of Kala–azar cases will eventually result in death (6; 5). In recent years, leishmaniasis outbreaks have been described with increasing frequency, including those in sub-tropical regions or regions not previously endemic across the global. In Brazil, beginning in 2005 (7), There is reports outbreaks of VL in different parts of the world like Nepal from 2004 – 2007 (8), China in 2014 (9), Kenya in 2008, 2011, 2013 and 2014 (10), Ethiopia in 2007 (11) different site with different number of cases.

VL is considered among the most neglected tropical diseases (NTD), is one of several emerging diseases of major public health importance in Ethiopia. An estimated 3.2 million people are at risk of VL in Ethiopia and VL is endemic in six regions of the country (2; 12). Based on the report of Oromia Regional Health Bureau NTD report; it is endemic in Borena, Guji and Bale Zones. Borena zone started reporting Leishmaniasis cases in 2012 from Arero, Dire and Miyo Districts. More cases are reported by Dire District and the problem expanded to additional districts and currently, cases are coming from six districts. The Zone Health Department (ZHD) reported relatively an increased number of suspected leishmaniasis cases in 2019, specifically from Magado Kebele.

Following reports of increased number of leishmaniasis cases in the zone, a team consisting of different experts drawn from Federal, Regional and WHO is established and deployed to the zone to provide technical support to ZHD from 14-25 October 2019. We conducted the investigation with a combination of epidemiological, entomological, and case control study and formed case definition by using a standard set of criteria to decide whether an individual should be classified as having the disease or not in this investigation. We further divided into two sub-teams each taking responsibility for case management and surveillance. The case management team dealt with verifications of case diagnosis, management and capacity building of health workers in Ya’abal’o hospital.

## Materials and methods

### Case definition

A case definition is formed by using a standard set of criteria to verify and decide whether an individual should be classified as having the disease or not in this investigation. Usually includes four components:

- Clinical information about the disease
- Characteristics about the people who are affected
- Information regarding the location or place
- Specification of time during which the outbreak occurred

#### VL case definition

A person who presents with fever for more than two weeks and an enlarged spleen (splenomegaly) **AND/OR** Enlarged lymph nodes (lymphadenopathy) **OR** either loss of weight, anaemia or leukopenia; while living in a known VL endemic area or having travelled to an endemic area.

### Study area and period

The study was conducted in Borena Zone, Oromia Region. The area is bordered in North, West Guji Zone, in South bordering Kenya, in West, Somali Regional State and in the West with South Nations, Nationalities and People Regional State. The 2019 projected total population of the affected area was 219,809 and most of the residents of the zone were pastoralists and most Districts were low lands. Entomological and environmental parts of the study were conducted in Magado village in Dire District of Borena Zone, which is endemic for Malaria and other NTDs.

### Study design

Descriptive design followed by case control study was conducted in the affected area.

### Descriptive studies

Based on case definition, we reviewed patients’ record from the health facilities to verify the cases. We included socio demographic variables such as age, sex, travel history, to endemic areas, vector control program, sleeping area/place, location, data of onset, date health facility visit and clinical information such as symptoms and treatment outcomes.

### Analytical Epidemiology

The dependent variable for the study was verified VL cases while independent variable includes variables such as age, sex, marital status, educational status, occupation and economic characteristics. We used case control design with 95% CI, 80% power, 30.8% controls exposure, and 3.73 odds ratio and with ratio of 1:2. All 33 verified cases and 66 controls were included in the study. We collected all variables’ information from cases admitted to hospital discharged using structured questionnaire by interviewing the patients, caregivers and respective control group selected from the same age group, sex and village. There may be limitation of recall bias from those returned to their home. All 99 study subjects were residents of Borena Zone for at least 2 years at time of diagnosis (cases) or at time of enrolment (controls). We included all cases of VL which fulfils cases definition, admitted to Ya’abal’o Hospital and discharged from 13th, July to 13th October, 2019. We entered the data to Epi info and exported to SPSS to analyse predictors using the logistic regression at significant value of P<0.05. Variable those identified for eligibility of binary logistic regression model at p-vale of 0.05 were again tested for final model of multivariate logistic regression analysis.

### Entomological and Environmental Study

We carried out collection of sand flies by using two CDC light trap and six sticky paper collection method for both indoor and outdoor to assess the biting and resting behavior of sand fly, species identification and parasite detection for environmental and entomological assessment. We also tried to identify environmental risk factors like availability of water sources, marshy areas, housing conditions, type of trees and domestic and/or wild animals by our visitation to Magado village in Dire district of Borena Zone. A total of 70 entomological specimens were collected. Out of the 70 specimens, 49 (70%) were collected from outdoor near house, 15 (21%) from bushes sites. Only 6 (9%) of the specimens were collected from indoors. Though the sample was not adequate for justification, the density is high in outdoor near to house compound. All mounted sand fly specimens was used to identify vector species based on identification key. However, PCR analysis for parasite detection could was not conducted due to absence of necessary reagent (Leishmania primers).

### Operational definitions

**Cases:** all verified VL cases admitted to Ya’abal’o hospital during the study period

**Control**: study participants having similar age group, sex and village with cases.

**Child dependent age group**: are participants of the study with age group ≤14 years old.

**Productive age group**: are participants of the study with age group from15-64 years old.

**Aged dependent age group:** are participants of the study with age group ≥65 years old.

## Results

### Descriptive report

#### Socio-demographic and economic characteristics

All participants (100%) were responded to our interview. Among 99 study subjects 33 (33.3%) of them were cases and 66 (66.7%) of them were controls. During interview, 21 (21.2%) respondents were treated cases and 78 (78.8%) of respondents were under treatment. Of the 99 participants 93 (93.9%) were male, 41 (41.4%) were have below the average family size of the country (4.8). Among all participants 5 (5%) were in aged dependent age group, 26 (26.3%) were in child dependent age group, and 68 (68.7%) participants were in productive age group. Of all participants 34 of them can’t read and write and 65 of them had different level of education. Among the participants 26 of them have no land; this is related to occupation i.e. pastoralists accounts 74 (74.7%), farmers 19 (19.2%) and others 6 (6.1%). The walling materials of their house were earth (mud) 86 (86.9%), wood 10 (10.1%) and thatched and brick 3 (3%) (Table 1).

**Table 1:**
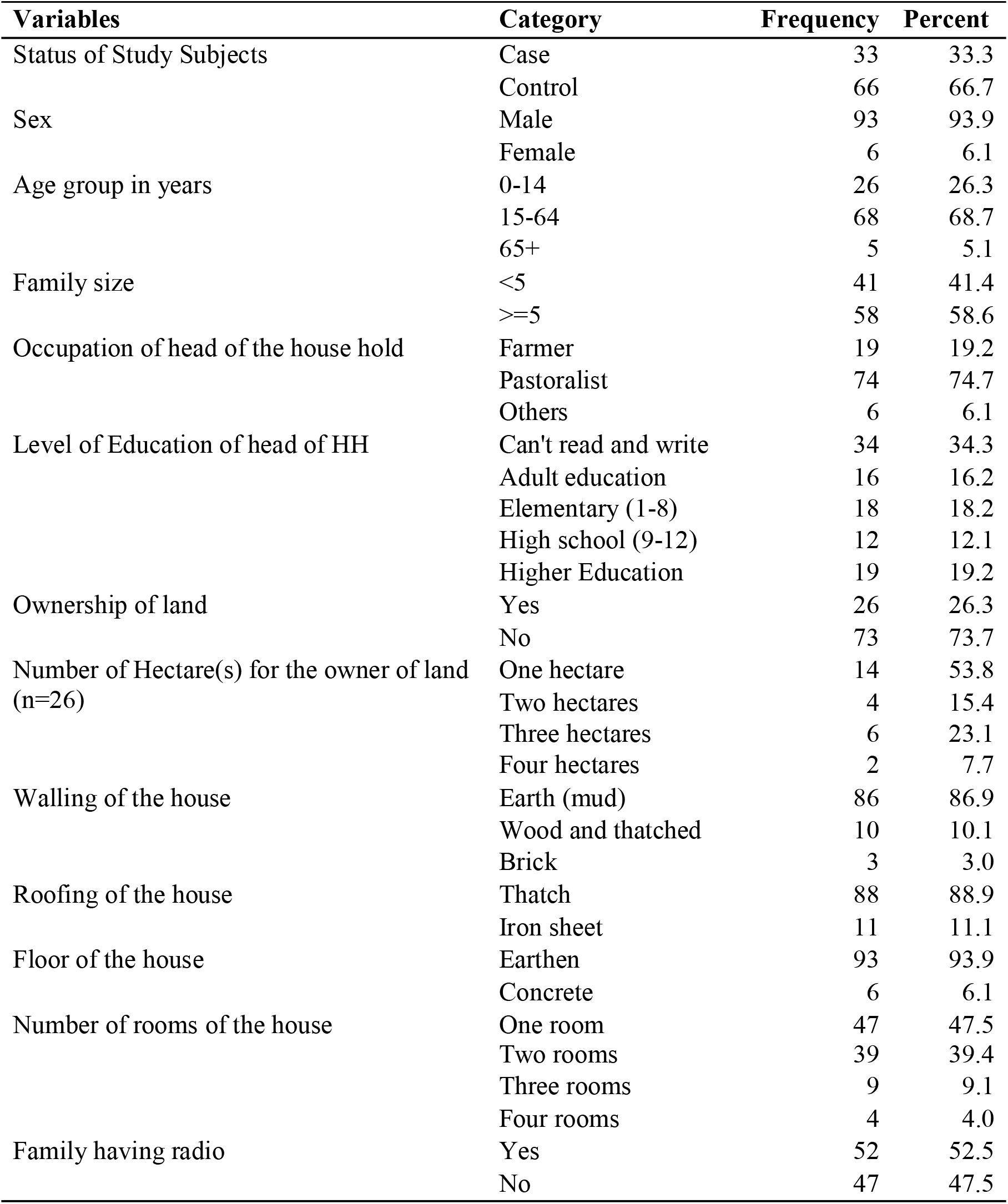
Socio-demographic Characteristics of Borena Zone, Oromia, Ethiopia 2019.

| **Variables** | **Category** | **Frequency** | **Percent** |
| --- | --- | --- | --- |
| Status of Study Subjects | Case | 33 | 33.3 |
|  | Control | 66 | 66.7 |
| Sex | Male | 93 | 93.9 |
|  | Female | 6 | 6.1 |
| Age group in years | 0-14 | 26 | 26.3 |
|  | 15-64 | 68 | 68.7 |
|  | 65+ | 5 | 5.1 |
| Family size | <5 | 41 | 41.4 |
|  | >=5 | 58 | 58.6 |
| Occupation of head of the house hold | Farmer | 19 | 19.2 |
|  | Pastoralist | 74 | 74.7 |
|  | Others | 6 | 6.1 |
| Level of Education of head of HH | Can't read and write | 34 | 34.3 |
|  | Adult education | 16 | 16.2 |
|  | Elementary (1-8) | 18 | 18.2 |
|  | High school (9-12) | 12 | 12.1 |
|  | Higher Education | 19 | 19.2 |
| Ownership of land | Yes | 26 | 26.3 |
|  | No | 73 | 73.7 |
| Number of Hectare(s) for the owner of land (n=26) | One hectare | 14 | 53.8 |
|  | Two hectares | 4 | 15.4 |
|  | Three hectares | 6 | 23.1 |
|  | Four hectares | 2 | 7.7 |
| Walling of the house | Earth (mud) | 86 | 86.9 |
|  | Wood and thatched | 10 | 10.1 |
|  | Brick | 3 | 3.0 |
| Roofing of the house | Thatch | 88 | 88.9 |
|  | Iron sheet | 11 | 11.1 |
| Floor of the house | Earthen | 93 | 93.9 |
|  | Concrete | 6 | 6.1 |
| Number of rooms of the house | One room | 47 | 47.5 |
|  | Two rooms | 39 | 39.4 |
|  | Three rooms | 9 | 9.1 |
|  | Four rooms | 4 | 4.0 |
| Family having radio | Yes | 52 | 52.5 |
|  | No | 47 | 47.5 |

#### Behavioural and environmental characteristics

Among all study participants 94 (94.9%) of them have no travel histories to other VL endemic area within the last 2 years, 76 (76.8%) HH were sprayed with anti-mosquito chemicals. Among 23 sprayed HHs 19 (82.6%) of them were sprayed ≤1 year duration. Among study participants 64 (64.6%) households had no bed net, 27 (77.1%) of them used always, 7 (20%) of them used sometimes & 1 household never used the net. Most of the time; 67 (67.8%) study subjects sleep inside the house and 32 (32.3%) of them sleep outside the house during night time. During night time 78 (78.8%) study subjects were sleep under acacia tree and during day time 95 (96%) of the have behaviour of sleeping under acacia tree (Table 2).

**Table 2:** Behavioural Characteristics in Borena Zone, Oromia, Ethiopia 2019.

| Variables | Category | Frequency | Percent |
| --- | --- | --- | --- |
| Travel history of study subjects to other VL endemic area | No | 94 | 94.9 |
|  | Yes | 5 | 5.1 |
| HH IRS status | No | 76 | 76.8 |
|  | Yes | 23 | 23.2 |
| Ownership status of dogs | No | 68 | 68.7 |
|  | Yes | 31 | 31.3 |
| Availability of 'Osole' near residential area | Yes | 80 | 80.8 |
|  | No | 19 | 19.2 |
| Family having Bed nets | Yes | 35 | 35.4 |
|  | No | 64 | 64.6 |
| Utilization of Bed nets during dry season (n=35) | Always used | 27 | 77.1 |
|  | Sometimes used | 7 | 20.0 |
|  | Never used | 1 | 2.9 |
| Utilization of Bed nets during rainy season (n=35) | Always used | 26 | 74.3 |
|  | Sometimes used | 8 | 22.9 |
|  | Never used | 1 | 2.9 |
| Sleeping place of study subjects | Inside room in the house | 67 | 67.7 |
|  | Outside of the house | 32 | 32.3 |
| Sleeping under acacia tree during night time | Yes | 78 | 78.8 |
|  | No | 21 | 21.2 |
| Sleeping under acacia tree during day time | Yes | 95 | 96.0 |
|  | No | 4 | 4.0 |

#### Visceral Leishmaniasis (VL) Morbidity and mortality

A total of 153 cases, of which 33 (22%) were verified for VL admitted in Ya’abal’o Hospital during July 2, 2019 to October 23, 2019. In addition, the hospital reported 9 deaths due to VL and 3 of them were verified for VL death. The AR and CFR among verified cases was 15/100,000 and 9.1% respectively.

#### Distribution of visceral leishmaniasis by time

Among the total 33 cases 15 (45.5%) of cases were reported with the date of onset in August 2019. The outbreak was started in June 20^th^, 2019 (Epi-week 25) increased gradually and reached its pick 5 cases in August 2019 (Epi-week 31) and showed decline ever week. There were no cases with the onset dates reported in Epi-week 39 and 40. Upon comparison of trends of cases reported from 2016 to 2019 there is a marked case surge in July and August 2019 more than other years of respective months (Fig 1-3).

**Fig 1.**
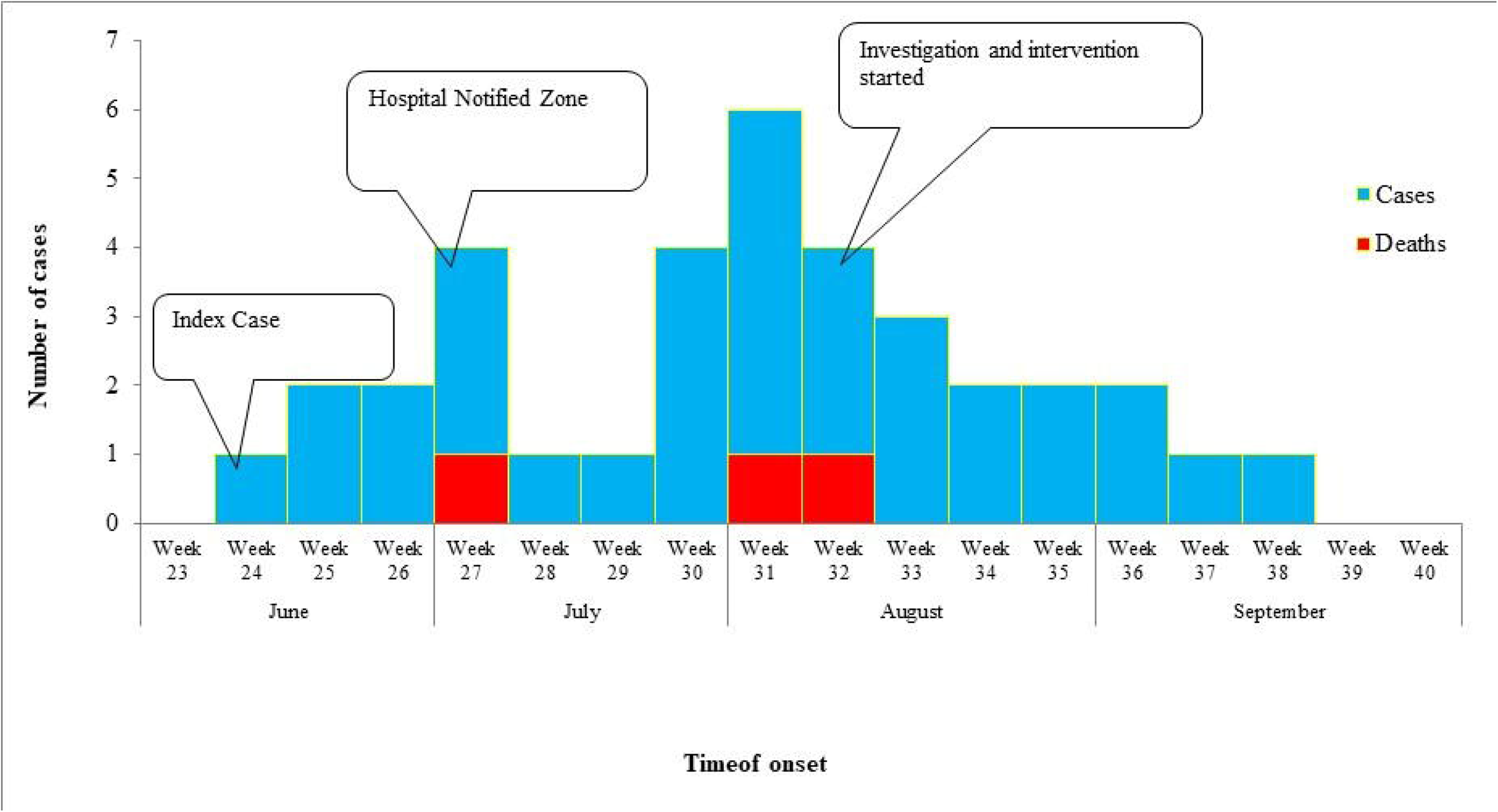
Epi Curve of suspected VL outbreak in Borena Zone, Oromia, Ethiopia 2019.

**Fig 2.**
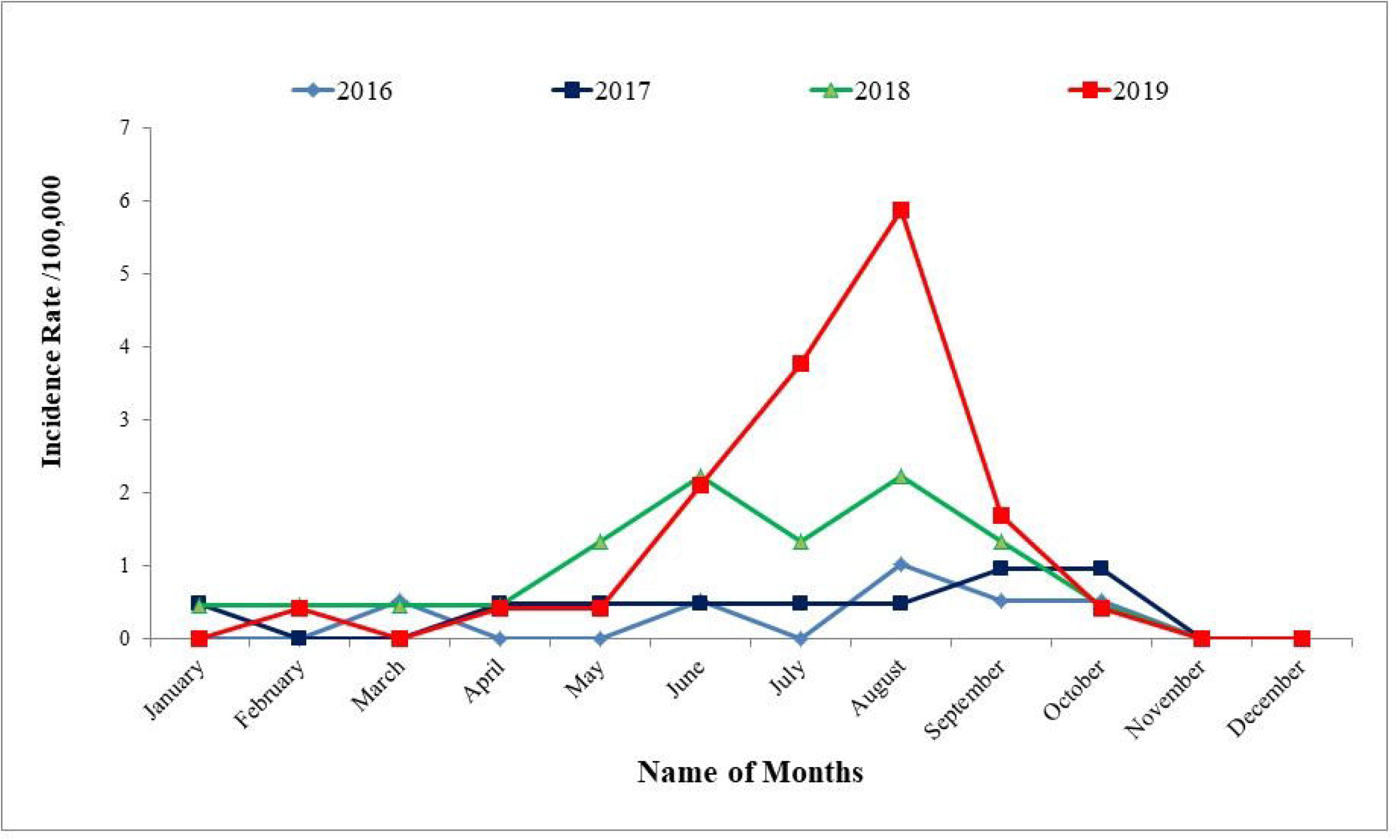
Monthly trend of VL cases in Borena 2016-2019, Oromia, Ethiopia.

**Fig 3.**
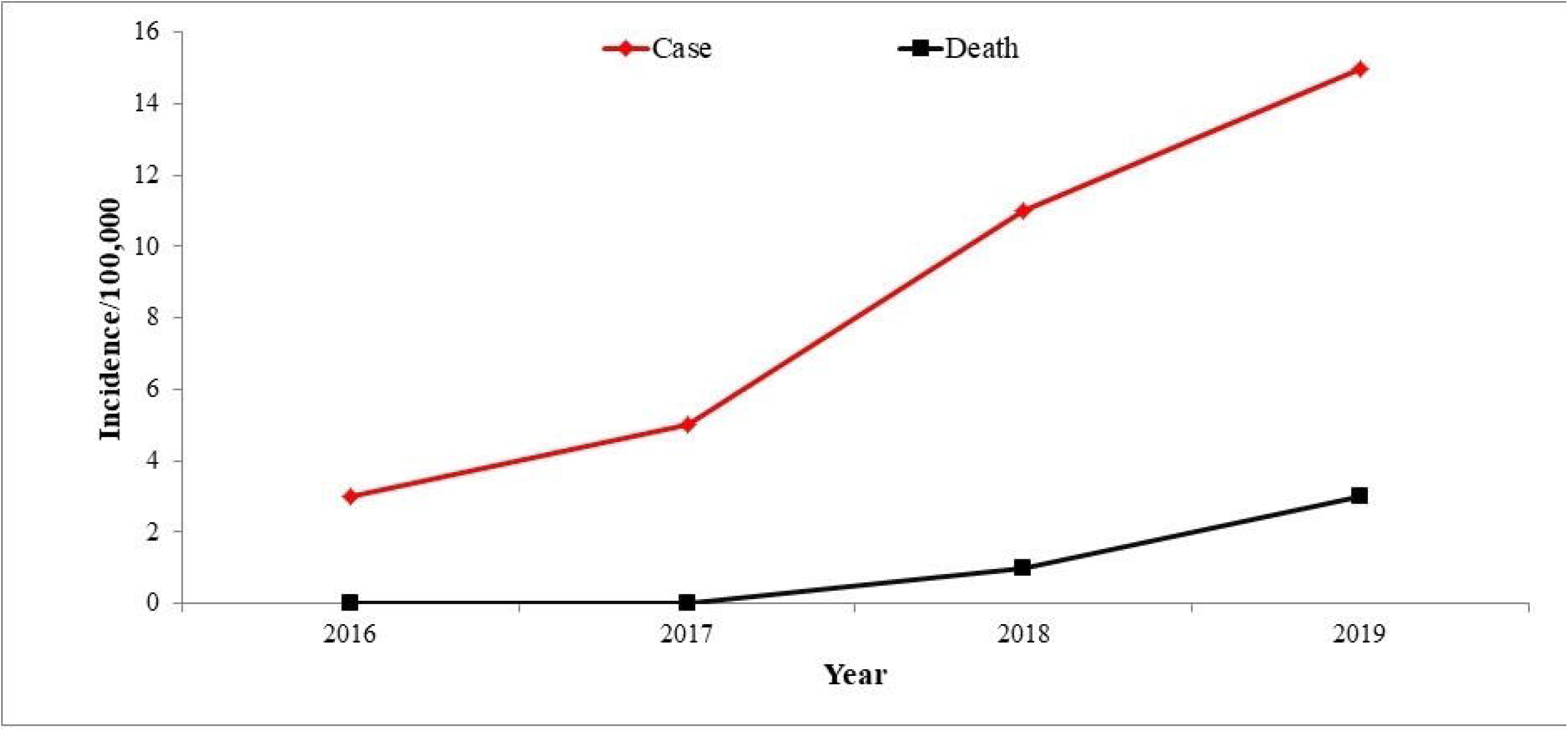
Annual trend of VL cases & deaths in Borena 2016-2019, Oromia, Ethiopia.

#### Distribution of visceral leishmaniasis by place

The AR among the Districts per 100,000 populations was: 51.2 in Dire, 6.6 in Elwaya, 6.2 in Dilo, 5.2 in Moyale, 3.3 in Dubuluk and 1.8 in Miyo districts. All confirmed deaths were from Dire (Table 3).

**Table 3.** AR and CFR of VL by District/100,000 populations Borena, Oromia, Ethiopia.

| Name of District | Total Population | Number of Cases | Number of Deaths | AR per 100,000 | CFR |
| --- | --- | --- | --- | --- | --- |
| Dire | 50,760 | 26 | 3 | 51.2 | 11.5 |
| Elwaya | 30,221 | 2 | 0 | 6.6 | 0 |
| Moyale | 37,991 | 2 | 0 | 5.3 | 0 |
| Dilo | 16,121 | 1 | 0 | 6.2 | 0 |
| Dubuluk | 30,285 | 1 | 0 | 3.3 | 0 |
| Mi'o | 54,431 | 1 | 0 | 1.8 | 0 |
| Total | 219,809 | 33 | 3 | 15 | 9.1 |

Among all cases; 26(79%) of them were from Dire District and Magado Kebele contributed 20 (77%) cases for Dire. Other 7 (21%) cases were from Moyale 2 cases, Elweya 2 cases, Dilo 1 case, Dubuluk 1 case and Miyo 1 case (Fig 4 and 5). More over all the cases from other Districts had travel history to Ele-Bora water point. All the confirmed three deaths were from Dire District.

**Fig 4.**
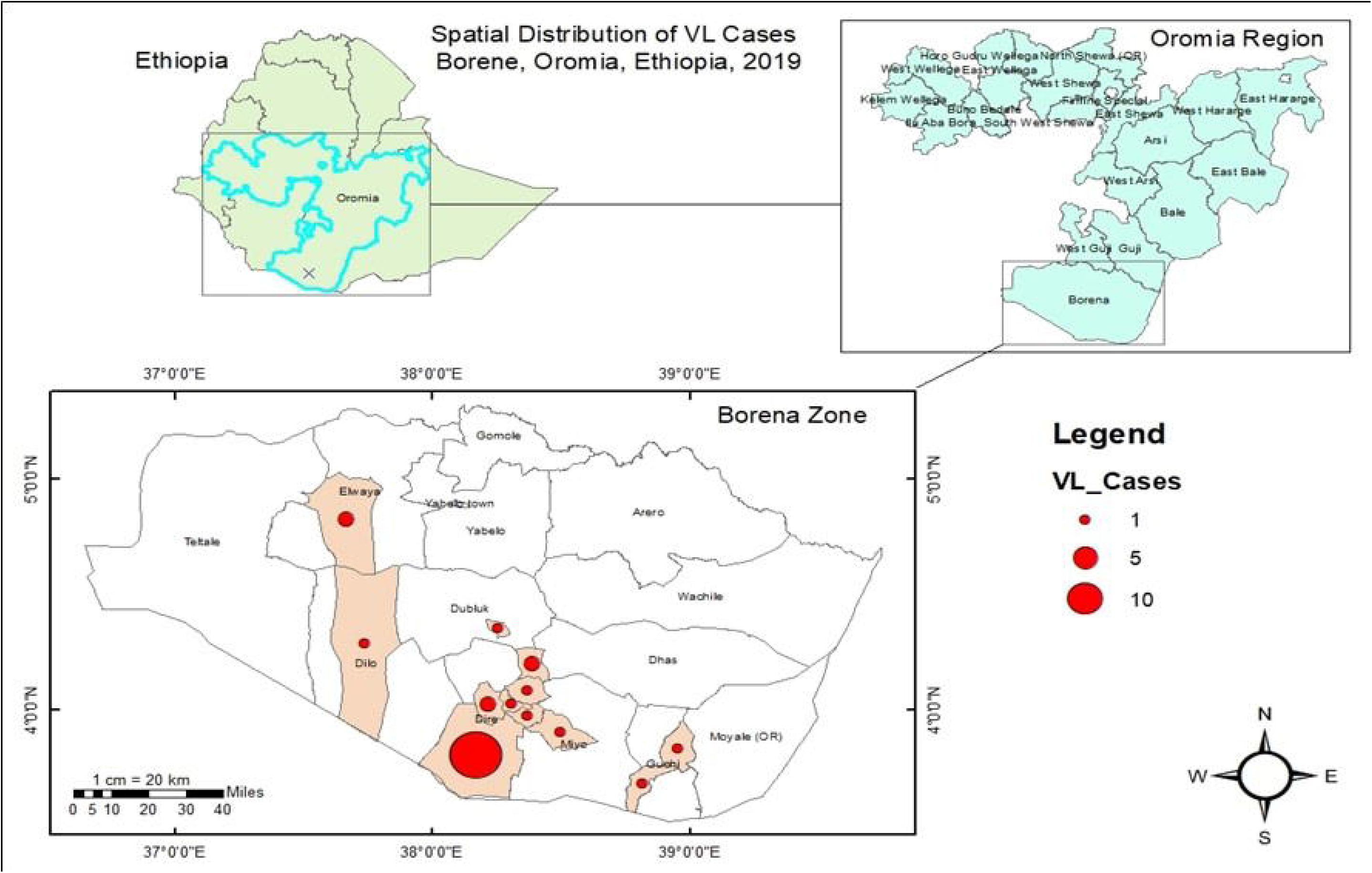
Spatial distribution of VL in Borena, Oromia, Ethiopia 2019.

**Fig 5.**
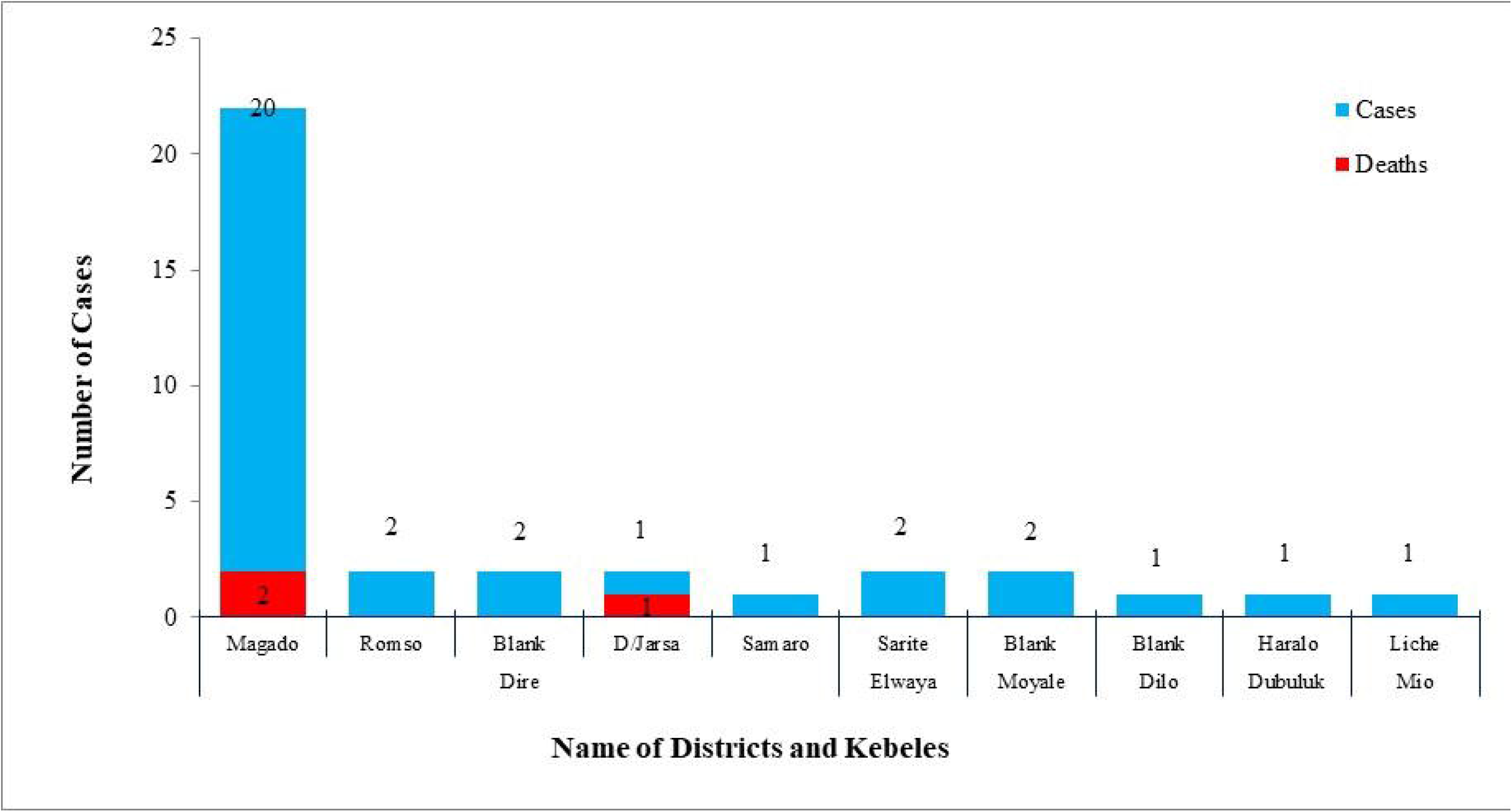
Number of VL cases by Districts in Borena Zone, Oromia Ethiopia 2019.

#### Distribution of visceral leishmaniasis by person

The AR per 100,000 populations with age groups was 105, 19.3 and 9.0 in 0-14 years, 15-64 years and 65+ years respectively. The age groups highly affected were 15-64 years with AR of 19.3/100, 000 population, while the CFR is high among age groups 0-14 years with CFR of 20%. This is may be due low immunity during child ood (Table 40 and Figure 59). Among all case 31 (94%) of them were male. There were only 2 female cases and both of them were alive.

**Table 4.** AR and CFR of VL with age group in Borena Zone, Oromia, Ethiopia, 2019.

| Age Category | Total population | Number of Cases | Number of Deaths | AR per 100,000 pop | CFR per 100 cases |
| --- | --- | --- | --- | --- | --- |
| 0-14 years | 94,936 | 10 | 2 | 10.5 | 20 |
| 15-64 years | 11,3743 | 22 | 1 | 19.3 | 4.5 |
| 65+ years | 11,130 | 1 | 0 | 9.0 | 0.0 |
| Total | 219,809 | 33 | 3 | 15.0 | 9.1 |

### Analytic Epidemiology findings

Study participants with adult education are 30 times more likely to have VL than those of having higher educational level (95% CI of AOR=2.378, 389.602), and similarly primary level of education are 13 times more likely to develop VL than those of having higher educational level (95% CI of AOR=1.107, 168.565). HH heads not able to read and write 93% less likely to be free from VL than those able to read and write (95% CI of AOR=0.007,0.582) and not owning land is 72% less likely to be free from VL than those owning land (95% CI of AOR=0.078,0.996). Participants not having bed-nets are 9 times more likely to be infected with VL than those having bed-nets (95% CI of AOR=1.763, 46.205). Living in house with walling of brick is 95% less likely to be susceptible for VL infection than those living in house with walling of earth (mud)(Table 5).

**Table 5.** Multivariate Analysis of VL risk factors in Borena Zone, Oromia, South Ethiopia.

| Risk Factors | Categories | Cases |  | Controls |  | COR (95%CI) | AOR (95% C.I.) | P-Value |
| --- | --- | --- | --- | --- | --- | --- | --- | --- |
|  |  | Number | Percent | Number | Percent |  |  |  |
| Level of Education of study subject | Can't read & write<sup>1</sup> | 16 | 47.1% | 18 | 52.9% |  |  | .055 |
|  | Adult education ** | 8 | 50% | 8 | 50% | 7.55 (1.51,37.89) | 30.44 (2.378, 38.60) | .009 |
|  | Primary (1-8) * | 6 | 33.3% | 12 | 66.7% | 8.50 (1.46,49.54) | 13.66 (1.10,168.56) | .041 |
|  | High school (9-12) | 1 | 8.3% | 11 | 91.7% | 4.25 (0.73,24.77) | 0.42 (0.01,26.08) | .686 |
|  | Higher education | 3 | 10.5% | 18 | 89.5% | 0.77 (0.62,9.58) |  |  |
| Ability of household head to read and write | No * | 17 | 26.2% | 48 | 73.8% | 0.398 (0.17,0.95) | 0.065 (0.007,0.582) | .015 |
|  | Yes<sup>1</sup> | 16 | 47.1% | 18 | 52.9% |  |  |  |
| Family ownership status of land | No* | 20 | 27.4% | 53 | 72.6% | 0.377 (0.15,0.95) | 0.278 (0.08,0.99) | .049 |
|  | Yes<sup>1</sup> | 13 | 50% | 13 | 50% |  |  |  |
| Having bed net | No** | 29 | 45.3% | 35 | 54.7% | 6.421 (2.03,20.32) | 9.024 (1.76,46.21) | .008 |
|  | Yes<sup>1</sup> | 4 | 11.4% | 31 | 88.6% |  |  |  |
| Walling of the house | Earth<sup>1</sup> | 26 | 30.2% | 60 | 69.8% |  |  | .092 |
|  | Brick* | 0 | 0% | 3 | 100% | 0.185 (0.05,0.78) | 0.052 (0.004,0.739) | .029 |
|  | Other | 7 | 70% | 3 | 30% | .000 | .000 | .999 |

Note:
\*\* is P-value < 0.01,
\* is P-value <0.05
<sup>1</sup> Reference

### Entomological findings

A total of 70 entomological specimens were collected. Out of the 70 specimens, 49 (70%) were collected from outdoor sites and 15 (21%) were from vegetation. Only 6 (9%) of the specimens were collected from indoors. Though the sample was not adequate for justification, the density is 8-10 flies per A4 size slide in outdoor near to house compound. In Ele-Bora village black cotton soil is important breeding site for sand fly, the community prefers to sleep outside house due to hot climatic condition results in an increased risk of infection. However, 9% sand fly was collected inside the houses, are not constructed well, have many holes in the wall, so it increases the risk of bite when a person sleeps even inside the house.

The descriptive part of our finding also shows among all study participants 68.7% of them own dogs and 80.8% of them respond that availability of ‘Osole’ near their residential area. Both dogs and ‘Osole’ are the reservoirs of sand flies. Additionally; acacia tree which is favorable for the breading of sand fly found in the area and 78.8% of the respondents sleep under this tree during night time.

### Environmental findings

Based on the data registered in Ya’abal’o hospital shows high cases were from Dire District specifically from Magado Kebele. We tried to identify environmental risk factors for VL in Magado Kebele. Accordingly; there was a deep well water source project in Ele-Bora village having tankers and ditches without water which makes the surrounding favourable area for breading of sand flies. For the sake of water source project the community obliged to come and dwell in the area for grazing and getting water for their cattle. Additionally; the area is marshy/swampy during rain; when heavy rain passes the land become cracked and makes small holes which helps for breading of sand flies and related insects.

The walling of house made of wood and thatched having many holes, roofing from thatched and plastic material, and floor from earth not easily cleanable having cracks and holes. There were also acacia trees in the district which serves as shading from sunlight during day time and as a shelter near herd of cattle for young male during night time. There were domestic animals like dogs and wild animals like “Osole” inhabit in the area

## Discussion

The increment in the number of cases in July and August 2019 might be by the awareness raising activities in the community by the zone health department and respective Districts which can improve the care seeking behaviour of the community and the attention given for complete data recording in the current suspected outbreak.

The number of cases is pitched in Magado Kebele of Dire District. The possible cause for increased number of cases in Magado might be related to settlement of community near water point of Ele_Bora village which is favourable for breading of Sand Flies which are responsible vector for Leishmania infection (3; 8). Additionally the entomological information collected from Ele_Bora village also indicates the presence of sand fly in the village. More over all the cases from other District had travel history to the water point in Dire District.

The age groups highly affected were 15-64 years male due to high risk of exposure to the breading sites of sand fly during outside work activities; in similar way study conducted in North Ethiopia also shows higher number of VL cases were recorded above 14 years of age group, and studies in Libo Kemem showed that males were affected more than females. In contrast to our study the study conducted in South Sudan shows 56% of the cases were under 5 years old (13). The reason for difference could be behavioural and cultural difference between the communities. The reason for similarity of the studies might be due to culture and habits of the male were engaged to keep cattle; they sleep at night time outside the house near their cattle and stays under shed of trees at day time (14). Domestic animals like dog accompany them to keep their cattle during day and night, which are the factors for the transmission of Leishmaniasis (15). The respondents also told that there are wild animals like “Osole” which are another risk factor for the transmission of the disease (16). The result shows that the fatality rate is relatively high among child dependent age group. This might be due low immunity of children than the productive age groups (3).

Statistical analysis model we used shows that level of education of study participants were significantly associated with VL cases, which is similar with study conducted in North Ethiopia (14). Similarly study conducted in North Ethiopia showed that educational level below grade five boosted VL odds (16; 17). In contrast to this there were no evidences shown in studies conducted in Nepal, South Sudan and Libo Kemkem of North West Ethiopia (8; 13; 14). The difference might be due to socio cultural and geographical location difference of study areas.

We found that there is a strong association between VL and poor housing condition like walling material built from mud (earth)and similar study conducted in Nepal with those living in a thatched houses without windows having 3–4 times higher odds of Kala-azar (8). In Spain also living in a detached house, were all strongly associated with the prevalence of asymptomatic infection (18). Cracked walls may be favourable area for the breading of and resting of sand flies and houses without window are free for the movement of vectors from outdoor to indoor flight. The study North Ethiopia also shows similar association (16).

Our study also indicates that owning specific land has significant association with VL, and similarly study conducted in Shebelle, Somali Region (12). Previous outbreaks were often related to force migration of non-immune populations into endemic areas following conflict (19). The reason behind might be due to high mobility of the community for grazing and not expected to construct well designed house which hinders the movement and breading of sand fly.

Bed-net is other predictor that is associated to VL cases. Similarly studies proved that having bed nets and utilization of bed nets have significant association with the prevalence of this morbidity (16; 17). This holds true that ITNs are protective factor for the sand flies mechanically as well as chemically by killing the vectors (20; 2). However, like IRS, the usefulness of Long Lasting ITNs very much depends on the biting behaviour of the vectors (indoor vs. outdoor). Another issue regarding the use of nets against sand flies is that much sand fly biting activity occurs during early evening between 19-21 o’clock before most people go to sleep so that exposure to sand fly bites is only reduced but not eliminated (2).

To conclude the findings: male productive age group were the affected group, level of education, ownership of land, having and utilization of bed nets and housing conditions are significantly associated with Visceral Leishmaniasis. Based on the main findings of our study we recommend that: formulating policies and guidelines on awareness creation for male productive age group regarding feeding habit of sand fly and prevention mechanisms control methods, Educating the community on prevention mechanisms like using repellents and safe sleeping mechanisms and additionally further investigation on the study area is the best remedy to overcome future VL outbreak occurrence.

## Data Availability

Raw data of the manuscript and line list of Visceral Leishmaniasis cases

## Acknowledgements

We would like to thank Addis Ababa University; College of Health Science; School of Public Health and Oromia Regional Health Bureau for the facilitation of investigation. Our greatest gratitude goes to outbreak investigation team for unrestricted technical contribution to overcome the outbreak status. Finally; our appreciation goes to Borena Zone Health Office, Ya’abal’o Hospital and their respective health workers as well as respondents for their willingness to avail requested data.

## Abbreviations

AOR: Adjusted Odds Ratio
AR: Attack Rate
CDC: Centre for Disease Prevention and Control
CFR: Case Fatality Rate
CI: Confidence Interval
COD: Crude Odds Ratio
FMoH: Federal Ministry of Health of Ethiopia
HH: Household
ITNs: Insecticide Treated Nets
NTD: Neglected Tropical Diseases
PHEM: Public Health Emergency Management
RHB: Regional Health Bureau
VL: Visceral Leishmaniasis
WHO: World Health Organization
ZHD: Zonal Health Department

## Annex

### List of figures

VL_Manuscript_Figures\Fig 1 Epi Curve of suspected VL outbreak in Borena Zone, Oromia, Ethiopia 2019.jpg VL_Manuscript_Figures\Fig 2 Monthly trend of VL cases in Borena 2016-2019, Oromia, Ethiopia.jpg VL_Manuscript_Figures\Fig 3 Annual trend of VL cases & deaths in Borena 2016-2019, Oromia, Ethiopia.jpg VL_Manuscript_Figures\Fig 4 Spatial distribution of VL in Borena, Oromia, Ethiopia 2019.jpg VL_Manuscript_Figures\Fig 5 Number of VL cases by Districts in Borena Zone, Oromia Ethiopia 2019.jpg VL_Manuscript_Figures\Fig 6 VL Distribution by age group in Borena, Oromia, Ethiopia 2019.jpg

**Fig 6.**
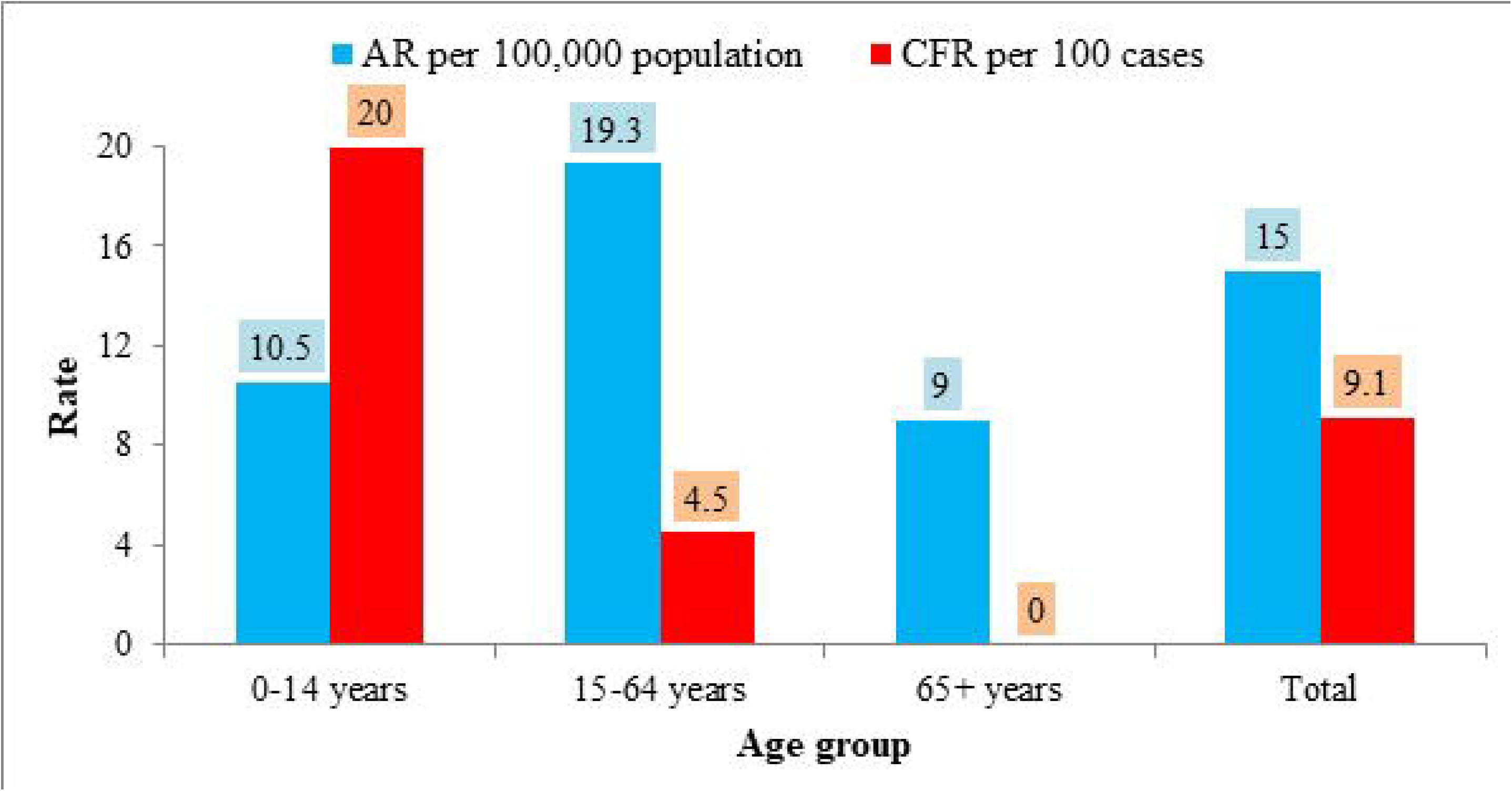
VL Distribution by age group in Borena, Oromia, Ethiopia 2019.

## References

1. Giradoni, Luigi. MANUAL ON CASE MANAGEMENT AND SURVEILLANCE OF THE LEISHMANIASES IN THE WHO EUROPEAN REGION. DK-2100 Copenhagen, Denmark: World Health Organization Regional Office for Europe, 2017. ISBN 978 92 89052 51 1.

2. FMoH. Guideline for Diagnosis, Treatment and Prevention of Leishmaniasis in Ethiopia 2nd Edition. Addis Ababa: FMoH, 2013.

3. Sundar, Shyam. Harrison’s Principles of internal Medicine 19th Edition. USA: McGraw-Hill Education, 2012. ISBN: 978-0-07-180216-1.

4. Epidemiological Investigation of Visceral Leishmaniasis Caused by Leishmania martiniquensis in a Non-endemic Area of Thailand. OSIR. 2, p. 1–7, Thailand: OSIR, June 2016, Vol. 9.

5. Ministry of health and population, Government of Nepal. National Guideline on Kala-azar Elimination Program (Updated). Teku, Kathmandu: Departyment of Health Service, Epidemiology and Disease Control Division, 2019.

6. Health, Ministry of. Guidelines for diagnosis, treatment and prevention of visceral leishmaniasis in South Sudan. Juba, South Sudan: Ministry of Health,.

7. Multilocus Sequence Analysis for Leishmania braziliensis Outbreak Investigation. Marlow MA, Boité MC, Ferreira GEM, Steindel M, Cupolillo E. 2, Berlin, Germany: Plos: Neglected Tropical Disease, 2014, Vol. 8. e2695. doi:10.1371/journal.pntd.0002695.

8. An outbreak investigation of visceral leishmaniasis among residents of Dharan town, eastern Nepal, evidence for urban transmission of Leishmania Donovani. Surendra Uranw, Epco Hasker. 21, Dharan: Article in BMC Infectious Diseases ·, January 2013, Vol. 13. DOI: 10.1186/1471-2334-13-21.

9. Investigation of outbreak of Visceral Leishmaniasis in 2014 in Jiashi County of Xinjiang. OSMAN Yisilayin, SIMAYI Adili. 5, Xinjiang: Chin J Parasitol Parasit Dis, 2015, Vol. 33. 1000-7423(2015)-05-0357-05.

10. WHO. PREVENTION, DIAGNOSIS AND TREATMENT OF VISCERAL LEISHMANIASIS (KALA-AZAR) IN KENYA National guidelines for health workers. Nairobi: REPUBLIC OF KENYA, MINISTRY OF HEALTH, 2017.

11. Kala-Azar Outbreak in Libo Kemkem, Ethiopia: Epidemiologic and Parasitologic Assessment. Jorge Alvar, * Seife Bashaye, Daniel Argaw, Israel Cruz. (2), pp. 275–282, Geneva: The American Society of Tropical Medicine and Hygiene, 2007, Vol. 77.

12. Epidemiology of visceral leishmaniasis in Shebelle Zone of Somali Region, eastern Ethiopia. Getachew Alebie*, Amha Worku, Siele Yohannes,. 209, Jigjiga: BMC: Parasites & Vectors, 2019, Vol. 12. https://doi.org/10.1186/s13071-019-3452-5.

13. Risk factors for the transmission of kala-azar in Fangak, South Sudan. Nyunguraa JL, Nyambatib VCS, Muitac M and Eric Muchirid E. pp 26–29, South Sudan: SSMJ, 2011, Vol. 4.

14. Visceral Leishmaniasis and Associated Risk Factors in Libo Kemkem,. Walelign Azene, Sissay Menkir, Ameha Kebede and Fikru Gashaw. 5, Northwestern Ethiop: EC MICROBIOLOGY, 2017, Vol. 7. pp 162–172.

15. Visceral Leishmaniasis in Ethiopia: An Evolving Disease. Samson Leta, Thi Ha Thanh Dao, Frehiwot Mesele and Gezahegn Alemayehu. 9, Adami Tullu, Ziway, Ethiopia: PLOS Neglected Tropical Diseases,2014, Vol. 8. e3131.

16. Risk factors of visceral leishmaniasis: a case control study in north-western Ethiopia. Solomon Yared, Kebede Deribe and Araya Gebreselassie. 470, Nort Ethiopia: Parasites & Vectors, 2014, Vol. 7. http://www.parasitesandvectors.com/content/7/1/470.

17. Preliminary survey of domestic animal visceral leishmaniasis and risk factors in north-west Ethiopia. Ambaye Kenubih, Shimelis Dagnachew and Gizat Almaw. 2 pp 205-210, Bishoftu Ethiopia: Tropical Medicine and International Health, 2015, Vol. 20. doi:10.1111/tmi.12418.

18. Revalence of asymptomatic Leishmania infection and associated risk factors, afer an outbreak. Ana Victoria Ibarra-Meneses, Eugenia Carrillo and Javier Nieto. 22, South Western Madrid, Spain: Euro Surveill, 2019, Vol. 24. https://doi.org/10.2807/1560-7917.ES.2019.24.22.1800379.

19. Leishmaniasis. Sakib Burza, Simon L Croft, Marleen Boelaert. 951-70, s.l.: The Lancet, 2018, Vol. 392. http://dx.doi.org/10.1016/.

20. Consortium, Malaria. LEISHMANIASIS CONTROL IN EASTERN AFRICA: PAST AND PRESENT EFFORTS AND FUTURE NEEDS: Situation and Gap Analysis. COMDIS:, University of Leeds, UK: Malaria Consortium, 2010.

